# Blood Gene Expression Risk Profiles and Interstitial Lung Abnormalities: COPDGene and ECLIPSE cohort studies

**DOI:** 10.1101/2022.01.31.22270173

**Authors:** Matthew Moll, Brian D. Hobbs, Aravind Menon, Auyon J. Ghosh, Rachel K. Putman, Takuya Hino, Akinori Hata, Edwin K. Silverman, John Quackenbush, Peter J. Castaldi, Craig P. Hersh, Michael J. McGeachie, Don D. Sin, Ruth Tal-Singer, Mizuki Nishino, Hiroto Hatabu, Gary M. Hunninghake, Michael H. Cho

**Affiliations:** Channing Division for Network Medicine, Brigham and Women’s Hospital, Boston, MA 02115, USA; Division of Pulmonary and Critical Care Medicine, Brigham and Women’s Hospital, Boston, MA 02115, USA; Harvard Medical School, Boston, MA 02115, USA; Center for Pulmonary Functional Imaging, Department of Radiology, Brigham and Women’s Hospital, Boston, MA 02115, USA; Department of Biostatistics, Harvard T.H. Chan School of Public Health, Boston, MA 02115, USA; Division of General Internal Medicine and Primary Care, Department of Medicine, Brigham and Women’s Hospital, Boston MA, 02115; Centre for Heart Lung Innovation, St. Paul’s Hospital, and Department of Medicine (Respiratory Division), University of British Columbia, Vancouver, BC, Canada; COPD Foundation, Washington D.C., USA

## Abstract

**Rationale:** Interstitial lung abnormalities (ILA) are radiologic findings that may progress to idiopathic pulmonary fibrosis (IPF). Blood gene expression profiles can predict IPF mortality, but whether these same genes associate with ILA and ILA outcomes is unknown.

**Objectives:** To evaluate if a previously described blood gene expression profile associated with IPF mortality is associated with ILA and all-cause mortality.

**Methods:** In COPDGene and ECLIPSE study participants with visual scoring of ILA and gene expression data, we evaluated the association of a previously described IPF mortality score with ILA and mortality. We also trained a new ILA score, derived using genes from the IPF score, in a subset of COPDGene. We tested the association with ILA and mortality on the remainder of COPDGene and ECLIPSE.

**Measurements and Main Results:** In 1,469 COPDGene (training n=734; testing n=735) and 571 ECLIPSE participants, the IPF score was not associated with ILA or mortality. However, an ILA score derived from IPF score genes was associated with ILA (meta-analysis of test datasets OR 1.4 [95% CI: 1.2-1.6]) and mortality (HR 1.25 [95% CI: 1.12-1.41]). Six of the 11 genes in the ILA score had discordant directions of effects compared to the IPF score. The ILA score partially mediated the effects of age on mortality (11.8% proportion mediated).

**Conclusions:** An ILA gene expression score, derived from IPF mortality-associated genes, identified genes with concordant and discordant effects on IPF mortality and ILA. These results suggest shared, and unique biologic processes, amongst those with ILA, IPF, aging, and death.

**Key messages:** *What is the key question:* Interstitial lung abnormalities (ILA) are radiologic findings that may progress to idiopathic pulmonary fibrosis (IPF). Do blood gene expression profiles that predict IPF mortality also associate with ILA?

*What is the bottom line:* An ILA gene expression score, derived from IPF mortality-associated genes, was associated with ILA and all-cause mortality. This score identified genes with concordant and discordant effects on IPF mortality and ILA. Our results suggest shared, and unique biologic processes, amongst those with ILA, IPF, aging, and death.

*Why read on:* Our results lend insight into how gene expression profiles and biological pathways associated with IPF prognosis relate to ILA and all-cause mortality

## Introduction

Interstitial lung abnormalities (ILA) are specific radiologic findings detected on computed tomography (CT) scans^1–3^ and, in some instances, may represent, or progress to, pulmonary fibrosis^4,5^. Both ILA and idiopathic pulmonary fibrosis (IPF) are associated with pulmonary symptoms^2,6^, diminished lung function^1,2,4,5,7,8^, and mortality^9^. Despite these clinical similarities, genetic analyses reveal that these entities have both overlapping and distinct genetic risk alleles^10,11^. Taken together, the clinical and genetic evidence suggests that ILA and IPF possess both shared and unique pathobiology.

The risk of IPF and IPF mortality are not due to genetic variants alone^12^. Gene expression data can reflect the combination of both genetic variation and environmental factors that contribute to IPF pathogenesis (e.g. cigarette smoking)^13^. Herazo-Maya et al.^14^ used blood microarray data to develop a 52-gene IPF risk score that predicted mortality in multiple IPF cohorts^15^. Whether this IPF mortality risk score, or the specific genes in this risk score, are associated with ILA or ILA mortality in current and former smokers is not known. A peripheral blood signature that predicted ILA and mortality in ILA could be important for identifying early disease and those at risk for worse outcomes. In addition, shared gene expression features of IPF progression and ILA risk would highlight important biologic processes associated with the spectrum of interstitial lung disease from precursor lesions to irreversible fibrosis to death.

Therefore, we hypothesized that the IPF risk score would be associated with ILA and ILA-associated mortality, and that this risk would be driven by a subset of the genes in the IPF mortality risk score; this subset of genes may also lend insight into the biologic mechanisms relating ILA to fibrosis and death. To test this hypothesis, we utilized participants with visual ILA scoring and peripheral blood RNA-seq data from the Genetic Epidemiology of COPD (COPDGene) study^16^, and participants with peripheral blood microarray gene expression data from the Evaluation of COPD Longitudinally to Identify Predictive Surrogate End-points (ECLIPSE) study^17^. We first assessed the association of the previously developed IPF risk score with ILA and time-to-death. As we hypothesized that ILA risk would be captured by a subset of these IPF risk score genes, we also created a gene expression risk score, based on these same genes, optimized for the outcome of ILA. We assessed the performance and differences between the scores and used over-representation analyses to gain insight into the biologic processes most relevant to ILA and all-cause mortality.

## Methods

### Study populations

All study participants provided written informed consent. Each study center obtained institutional review board approval.

#### COPDGene

We included participants from the Genetic Epidemiology of COPD (COPDGene) study^16^ who had a 5-year follow up visit with blood RNA-sequencing (RNA-seq) data and computed tomography (CT) scans that were assessed for the presence ILA^9^. Details of the COPDGene study have been previously described^16^. Briefly, COPDGene is a prospective cohort study of non-Hispanic white (NHW) and African American (AA) smokers (≥10 pack-years of smoking), aged 45-80 years at study initiation, with and without COPD. The study was originally conceived of as a case-control study and has been extended into a longitudinal study with 5- and 10-year follow up visits. Whole blood samples, as well as anthropometric, spirometry, and CT imaging data were collected at each visit.

#### ECLIPSE

We included Evaluation of COPD Longitudinally to Identify Predictive Surrogate End-points (ECLIPSE) study^17^ participants with microarray gene expression data and chest CT scans that were assessed for the presence of ILA^9^. ECLIPSE participants were smokers (≥10 pack-years of smoking) aged 45-75 years at study enrollment. Baseline questionnaire, spirometry, CT imaging and blood samples were collected. In ECLIPSE, COPD participants, but not controls, were followed longitudinally for 3 years.

### ILA phenotyping

In both COPDGene and ECLIPSE, thoracic CT scans were assessed for ILA using a sequential method by three readers as previously described^10,11^. The definition of ILA included in this manuscript conforms to the updated definition utilized by the Fleischner society^3,18–20^.

### Preparation of gene expression data

#### RNA sequencing

COPDGene whole blood RNA-seq data was available at the 5-year follow up visit, and data generation was previously described^13^. Briefly, PAXgene Blood RNA tubes were used to collect whole blood samples, and the Qiagen PreAnalytiX PAXgene Blood miRNA Kit (Qiagen, Valencia, CA) was used to extract total RNA. Samples with concentrations > 25 ug/uL and RNA integrity number (RIN) > 6 were eligible for sequencing. TruSeq Stranded Total RNA with Ribo-Zero Globin kit (Illumina, Inc., San Diego, CA) were used for globin-reduction and cDNA library preparation. The Illumina HiSeq 2500 sequencer was used to generate 75 bp reads with a mean of 20 million reads per sample. Skewer^21^ was used to trim TruSeq adapter sequences.

Additional quality control was performed using FASTQC (https://www.bioinformatics.babraham.ac.uk/projects/fastqc/) and RNA-SeQC^22^. Reads were aligned to the human GRCh38 reference genome using STAR 2.5^23^. Count data were adjusted for library depth and batch effects were removed using the limma removeBatchEffects function^24^.

#### Microarray gene expression data

ECLIPSE whole blood microarray data were available at the initial enrollment visit. Details regarding microarray data collection and processing were previously published^25^. Total RNA was extracted using PAXgene Blood miRNA kits and hybridized to the Affymetrix Human Gene 1.1 ST array. Quality control metrics were implemented using the Bioconductor oligo^26^ and RMA Express^27^ packages. The Factor Analysis for Robust Microarray Summarization^28^ package was utilized for background correction and normalization. Batch effects were removed using the limma removeBatchEffects function^24^. As some gene transcripts were represented by multiple probes, we chose the probe with the greatest interquartile range, as previously described^14,15^.

We then took additional steps to facilitate comparability across RNA-seq and microarray data technologies: (1) we limited transcripts to those present in both data sets based on HGNC symbols; (2) we scaled and centered all gene expression data to have a mean of zero and a standard deviation of 1.

### Statistical analyses

#### Overview of study design

COPDGene was used as a discovery and testing cohort, and ECLIPSE was used for independent replication (Figure 1). In COPDGene, risk score training was performed in half the participants with the other half of the participants used for risk score testing. We assessed the association of three transcriptome-based risk scores (see *Predictors*) with outcomes (see *Outcomes*) in the COPDGene testing sample. We attempted to replicate the risk score associations in ECLIPSE.

**Figure 1:**
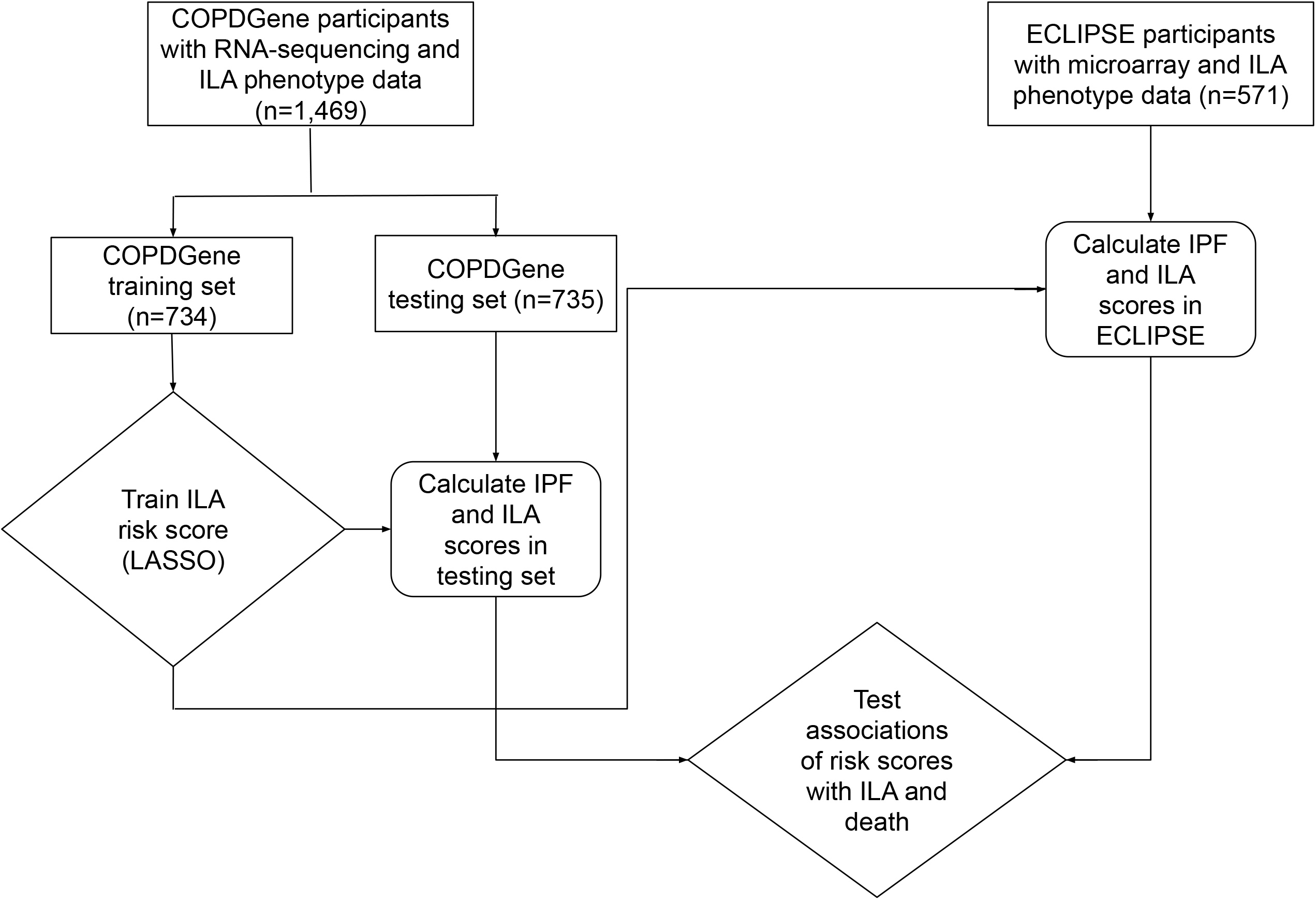
Schematic of study design. ILA = interstitial lung abnormalities. IPF = idiopathic pulmonary fibrosis. LASSO = least absolute shrinkage and selection operator.

#### Predictors

We calculated the Herazo-Maya et al.^14,15^ IPF prognosis signature using transcriptomic data from COPDGene and ECLIPSE and applied this risk signature to the prediction of ILA and mortality in these cohorts. Two transcripts were not available in our cohorts (*C2ORF27A* and *SNHG1*), resulting in 50 genes available for testing. Briefly, calculating this risk score involves calculating the proportion of up- and down-regulated genes, summing the normalized expression values, calculating the product between the summed normalized expression values and proportion of decreased and increased genes, and comparing the up- and down-scores to the median for the population. If both the up- and down-scores are higher than the median of the population in which the score is being calculated, then the person is considered to be at high IPF mortality risk (1= high IPF mortality risk, 0=low IPF mortality risk).

To test the performance of these same genes using an alternative method, we developed a risk score optimized to the outcome of ILA by taking the following steps:

1. We used the 50 available transcripts as inputs to construct a penalized regression (Least Absolute Shrinkage and Selection Operator (LASSO)) model in the COPDGene training set optimized to the outcome of ILA. LASSO regression shrinks coefficients toward zero to provide feature selection and minimizes collinearity amongst predictors. We tuned models within the COPDGene training set using 5-fold cross-validation, optimizing the area-under-the-receiver-operating-curve (AUC) on the left-out fold.
2. We used this penalized regression model (weights of gene transcripts) to calculate the log odds for ILA for each individual in the COPDGene testing sample and ECLIPSE.
3. We scaled and centered the risk scores within the COPDGene testing and ECLIPSE datasets separately.

Thus, we evaluated two main predictors in this study: (1) the original IPF mortality risk score, and (2) a re-weighted risk score optimized to ILA as an outcome (hereafter, “ILA score [IPF transcripts]”). Results were presented as 1 standard deviation increases in risk scores. Each risk score was tested in the COPDGene testing set and the ECLIPSE replication cohort. We additionally compared the direction of effects of genes in these scores. We range-standardized both risk scores and plotted histograms of predicted probabilities for each respective outcome in the COPDGene testing set.

As an additional analysis, we created an ILA-optimized risk score using genome-wide transcripts (hereafter, “ILA score [all transcripts]”). Prior to training the ILA score [all transcripts], we limited our analyses to highly expressed transcripts (>1 count per million in 99% of samples), resulting in 9,100 transcripts. These 9,100 transcripts were used as inputs into LASSO regression. The model was tuned within the COPDGene training set using 10-fold cross-validation, minimizing misclassification error on the left-out fold. We then calculated and standardized the risk scores above as for the ILA score [IPF transcripts]. We tested the association of each risk score with outcomes in the COPDGene testing set and ECLIPSE.

#### Outcomes

We examined two primary outcomes available in both cohorts: (1) prevalent ILA (at the time of blood sample collection), and (2) time-to-death. With respect to time-to-death, COPDGene participants were followed for up to 5 years after collection of RNA-seq data, and ECLIPSE participants for up to 8 years.

#### Models and model specifications

We used logistic regression to test the association of each of the 50 transcripts with ILA and compared the direction of effect for ILA to the transcript direction (up or down) in the IPF mortality score. We also compared the association of the 50 individual transcripts with all-cause mortality and compared the effect direction to both the effect for ILA association and direction in the original IPF mortality score. We assessed the association of each risk score (IPF score, ILA score [IPF transcripts], ILA score [all transcripts]) with ILA and time-to-death. Logistic regression was used to assess associations with ILA. We used Cox regression^29^ to test associations with time-to-death (survival R package^30^). Models were adjusted for age, sex, race, body-mass index, pack-years of cigarette smoking, and current smoking status (at the time of blood sample collection). For time-to-death analyses, we also performed stratified analyses in ILA and non-ILA participants. For ILA, the Bonferroni-adjusted threshold was 0.05/3 risk scores/2 cohorts = 0.0083. For time-to-death analyses, the Bonferroni-adjusted threshold was 0.05/3 risk scores/2 cohorts/3 strata = 0.0028. Proportional hazards assumptions were evaluated with Schoenfeld residual plots and tests. As a sensitivity analysis, we further adjusted models for white blood cell differential counts to assess whether white cell counts attenuated the observed signals. To combine the signals between the two testing cohorts (COPDGene testing, and ECLIPSE), we performed fixed-effects inverse variance-weighted meta-analyses using the meta R package^31^.

#### Mediation analyses

We noted that the associations of the ILA score [IPF transcripts] with time-to-death were substantially attenuated when age was added to models. Therefore, we performed causal mediation analyses to determine whether the effects of age on mortality were mediated through this score. We used the medflex R package^32^ to perform natural effects causal mediation analyses^33,34^ in the COPDGene testing set. We considered age as the exposure, and death (binary) as the outcome in logistic regression analyses. We considered the ILA score [IPF transcripts] as the mediator, and a p-value for the natural indirect effects less than 0.05 was considered significant.

#### Characterization of IPF and ILA score genes

To gain biologic insight into the relationship between genes that comprised the risk scores, we examined Pearson correlation coefficients between each of the 50 transcripts and constructed a heatmap of correlation coefficients. The observed correlation structure allowed us to use the sigora R package^35^ to perform pathway enrichment analyses for the 50 gene transcripts. We also evaluated how changing the number of genes in the ILA score [IPF transcripts] affected predictive performance (Supplementary Methods).

All analyses were performed in R version 4.0.3 (www.r-project.org). Normality for continuous variables was assessed by visual inspection of histograms. Results were reported as mean ± standard deviation or median [interquartile range], as appropriate. Continuous variables were compared with Student t-tests or Wilcoxon tests, and categorical variables were compared with analysis of variance (ANOVA) or Kruskal-Wallis tests, as appropriate. A p-value less than 0.05 was considered nominally significant, and p-values below Bonferroni-adjusted thresholds were considered significant.

## Results

### Characteristics of study participants

Figure 1 is a schematic of the study design. We included 1,469 COPDGene participants from the 5-year follow up visit with RNA-seq and visual scoring of ILA phenotype data, and 571 ECLIPSE participants with microarray and ILA phenotype data. Table 1 shows demographic characteristics and outcomes in the COPDGene training set (n=734), COPDGene testing set (n=735), and ECLIPSE (n=571). Characteristics were similar across COPDGene training and testing sets. Compared to COPDGene, ECLIPSE participants were more likely to be male, were all European ancestry, had more pack-years of smoking, were less likely to be current smokers, were less likely to have ILA, and had a higher proportion of deaths.

**Table 1:**
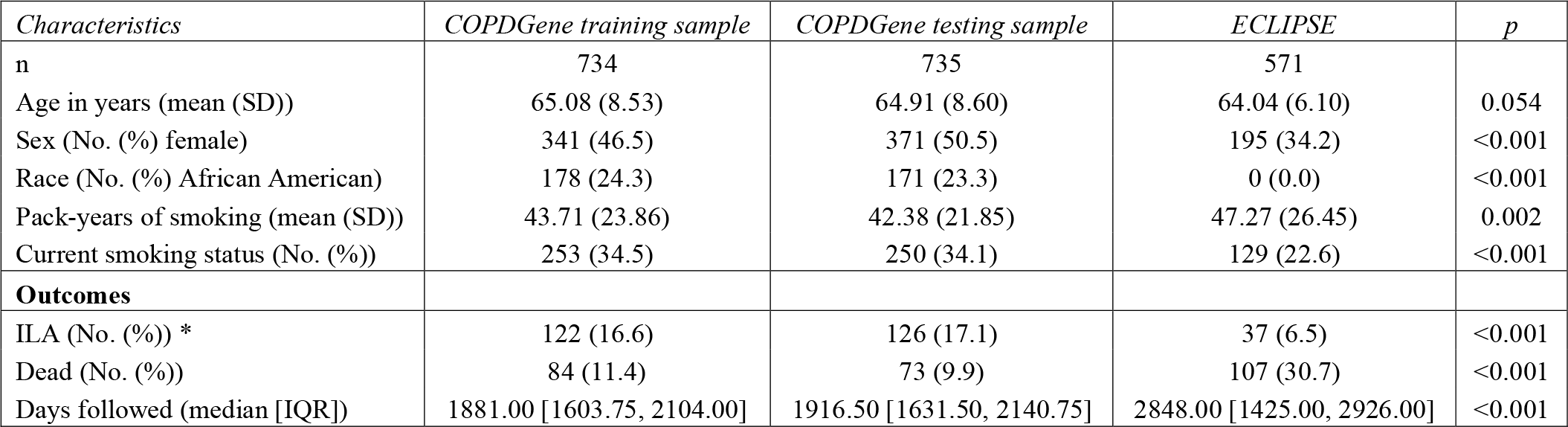
Characteristics of study participants. ILA=interstitial lung abnormalities. *ILA prevalence in the overall population was 5.7% (586/10,364) in COPDGene and 1.3% (37/2,746) in ECLIPSE.

| *Characteristics* | *COPDGene training sample* | *COPDGene testing sample* | *ECLIPSE* | *p* |
| --- | --- | --- | --- | --- |
| n | 734 | 735 | 571 |  |
| Age in years (mean (SD)) | 65.08 (8.53) | 64.91 (8.60) | 64.04 (6.10) | 0.054 |
| Sex (No. (%) female) | 341 (46.5) | 371 (50.5) | 195 (34.2) | <0.001 |
| Race (No. (%) African American) | 178 (24.3) | 171 (23.3) | 0 (0.0) | <0.001 |
| Pack-years of smoking (mean (SD)) | 43.71 (23.86) | 42.38 (21.85) | 47.27 (26.45) | 0.002 |
| Current smoking status (No. (%)) | 253 (34.5) | 250 (34.1) | 129 (22.6) | <0.001 |
| **Outcomes** |  |  |  |  |
| ILA (No. (%)) * | 122 (16.6) | 126 (17.1) | 37 (6.5) | <0.001 |
| Dead (No. (%)) | 84 (11.4) | 73 (9.9) | 107 (30.7) | <0.001 |
| Days followed (median [IQR]) | 1881.00 [1603.75, 2104.00] | 1916.50 [1631.50, 2140.75] | 2848.00 [1425.00, 2926.00] | <0.001 |

### Development of risk scores

Fifty out of the 52 genes from Herazo-Maya et al.^14,15^ were available in both cohorts (*C2ORF27A* and *SNHG1* were missing). First, using these 50 genes, we calculated the IPF score in COPDGene and ECLIPSE participants. We noted that when testing the associations of individual transcripts with ILA or death, 23 genes had a discordant direction of effect for ILA and 3 genes (*HLA-DP1, HLA-DP2, LPAR6*) had a discordant direction of effect for all-cause mortality compared to the directions of effects reported for the IPF score (Table E1). Second, we used the 50 available genes to construct a LASSO penalized regression model in the COPDGene training set, which was optimized to the outcome of ILA (ILA score [IPF transcripts]). Table E2 shows the ILA score [IPF transcripts] gene transcripts, beta coefficients and comparisons of directions of effects to the IPF risk score. Only three transcripts (*LBH, GBP4, BTN3A1*) were significantly associated with ILA, all of which were included in the ILA score [IPF transcripts]. The ILA score [IPF transcripts] included 11 transcripts (*BTN3A1, CPED1, CXCR6, GBP4, GPR174, IL7R, LBH, LPAR6, LRRC39, NAP1L2, PLBD1*), six of which had a discordant direction of effect with the IPF score (*BTN3A1, CPED1, GBP4, GPR174, LPAR6, NAP1L2*). The distribution of the ILA score [IPF transcripts] in both cohorts is shown in Figure E1. We then plotted range-standardized predicted probabilities in the COPDGene testing set and observed overlapping but distinct distributions between the two risk scores (Figure E2). We additionally used genome-wide transcripts to train a risk score to ILA (ILA score [all transcripts]); this score included 25 transcripts (Table E3), none of which overlapped with the 50 genes in the IPF score. We then tested these risk scores (IPF score, ILA score [IPF transcripts], ILA score [all transcripts]) for association with ILA and time-to-death in the COPDGene testing set and ECLIPSE.

### Association of risk scores with ILA

The IPF score was not associated with ILA in the COPDGene testing set or ECLIPSE (Table 2). The univariable associations of the ILA score [IPF transcripts] with ILA in both cohorts is shown in Figure 2A. In multivariable models (Table 2), the ILA score [IPF transcripts] was associated with ILA in the COPDGene testing set and ECLIPSE (meta-analysis OR 1.4 [95% CI: 1.2-1.6], p=6.8e-5). As a sensitivity analysis, we further adjusted models for white blood cell counts, and observed similar results (Table E4).

**Table 2:**
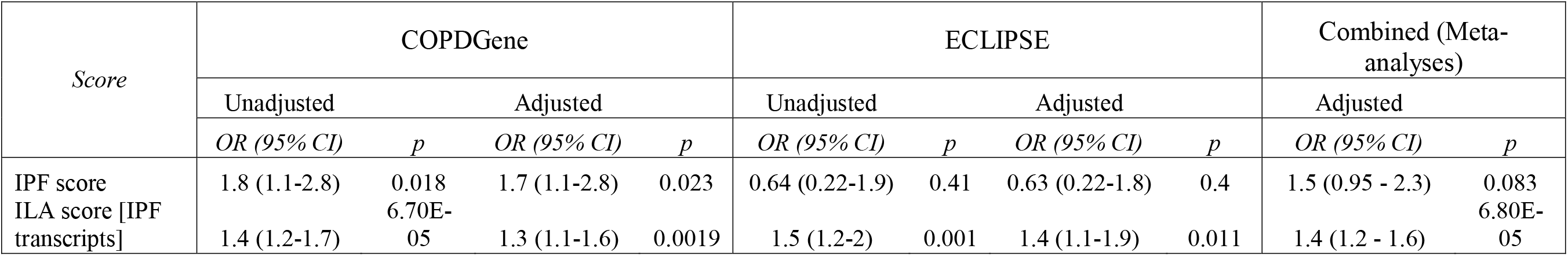
Odds ratios of the IPF and ILA [IPF transcripts] risk scores with ILA in the COPDGene test set (n=735) and ECLIPSE (n=571). Logistic regression models were adjusted for age, sex, race, body-mass index, pack-years of smoking, and current smoking status. Inverse variance fixed-effects meta-analyses of the adjusted estimates were performed. ILA=interstitial lung abnormalities. IPF=idiopathic pulmonary fibrosis.

**Figure 2:**
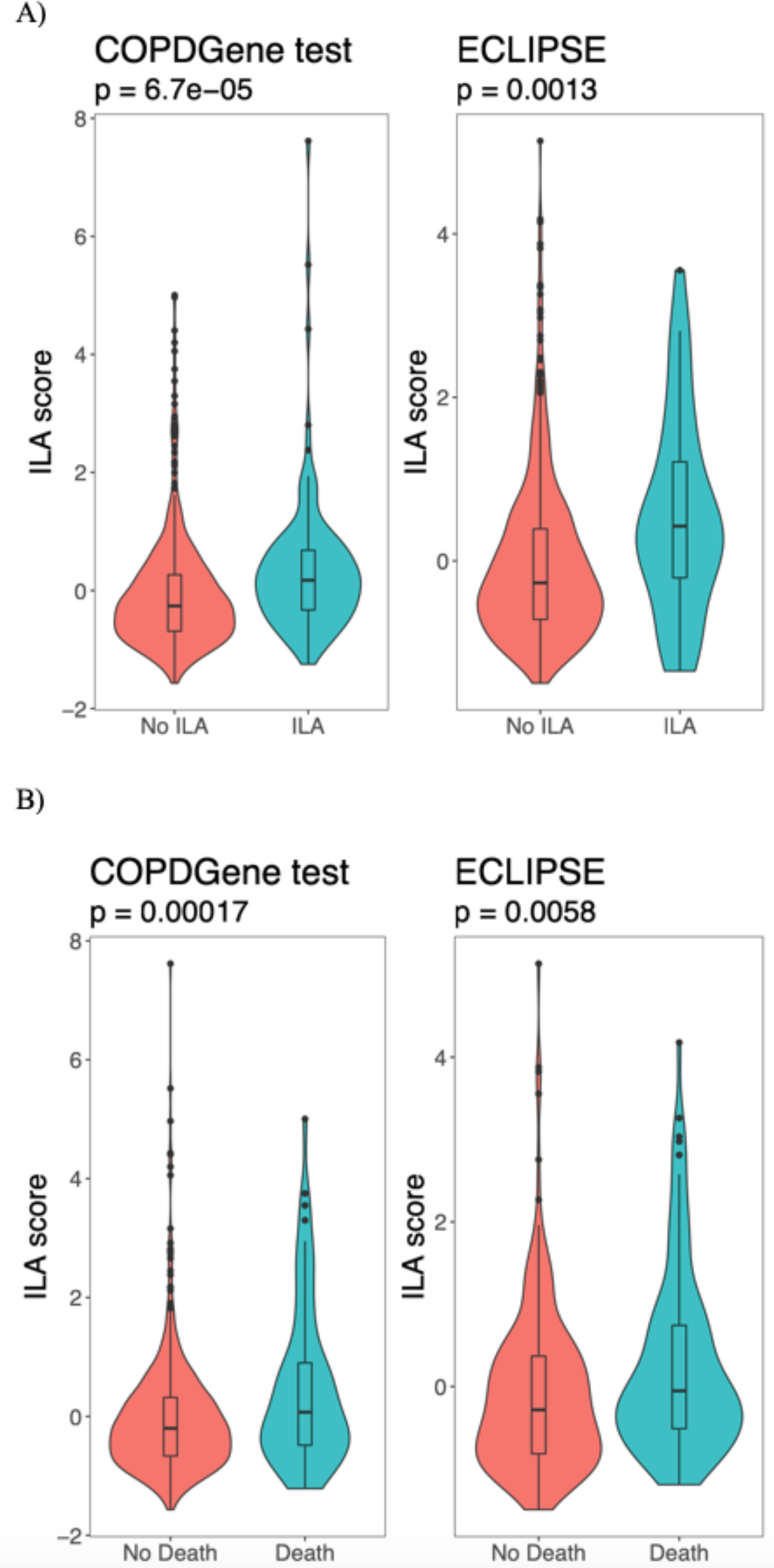
Violin and box plots showing the unadjusted association of the ILA score [IPF transcripts] with ILA (A) and death (B) in the COPDGene test set (n=735) and ECLIPSE (n=571).

### Association of risk scores with time-to-death

The IPF risk score was not associated with mortality in the COPDGene testing set or ECLIPSE (Table 3). The univariable associations of the ILA score [IPF transcripts] with death in both cohorts is shown in Figure 2B. In multivariable analyses (Table 3), the ILA score [IPF transcripts] is associated with time-to-death in all participants, but not the ILA participants. Meta-analyses demonstrate a significant association of the ILA score [IPF transcripts] with time-to-death (Figure 3) in all (meta-analysis HR 1.25 [95% CI: 1.12-1.41], p=1.25e-4) and non-ILA (meta-analysis HR 1.33 [95% CI: 1.17-1.52], p=2.17e-5) participants. As a sensitivity analysis, we further adjusted models for white blood cell counts and observed similar results (Table E5). In causal mediation analyses, we observed that the ILA score [IPF transcripts] mediated the effects of age on mortality (natural indirect effect p = 0.003; proportion mediated = 11.8% [95% CI: 4.04%-22.6%]).

**Table 3:**
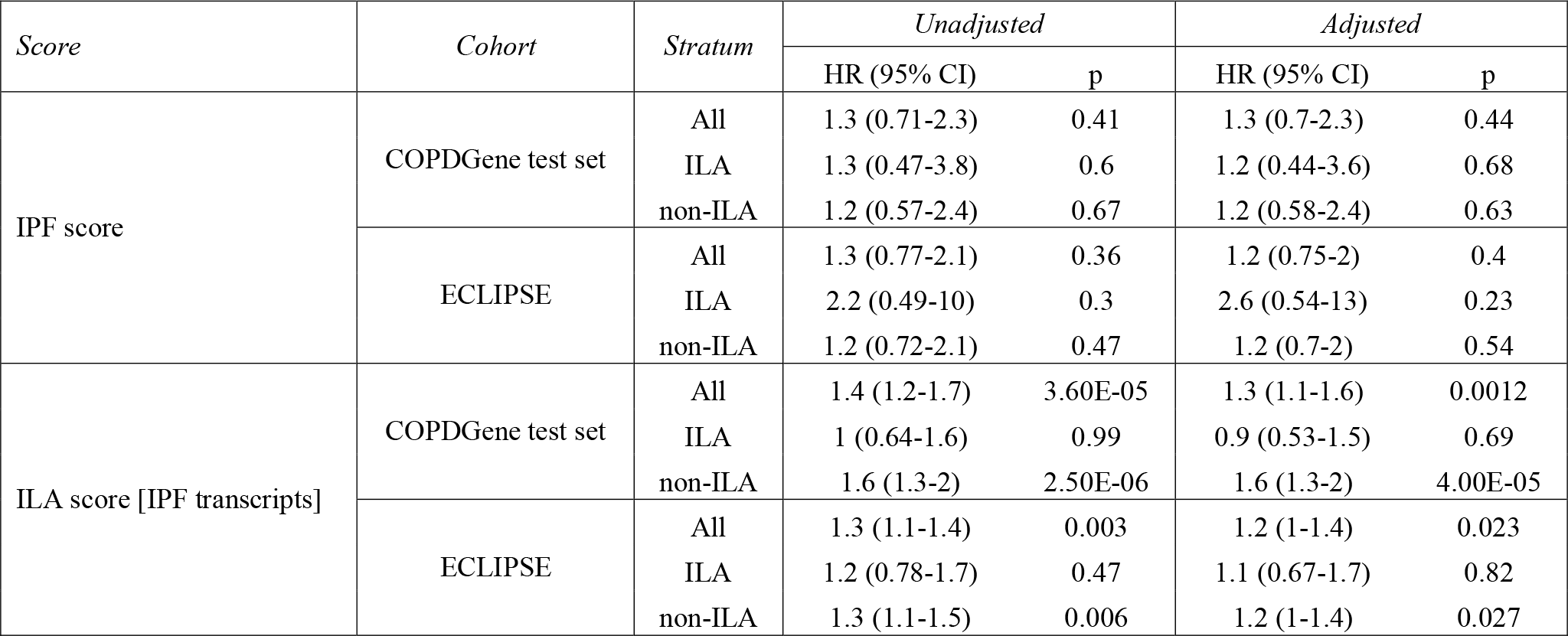
Association of the IPF and ILA [IPF transcripts] scores with time-to-death in the COPDGene test set (n=735) and ECLIPSE (n=571). Multivariable Cox regression models were adjusted for age, sex, race, body-mass index, pack-years of smoking, and current smoking status. ILA = interstitial lung abnormalities.

**Figure 3:**
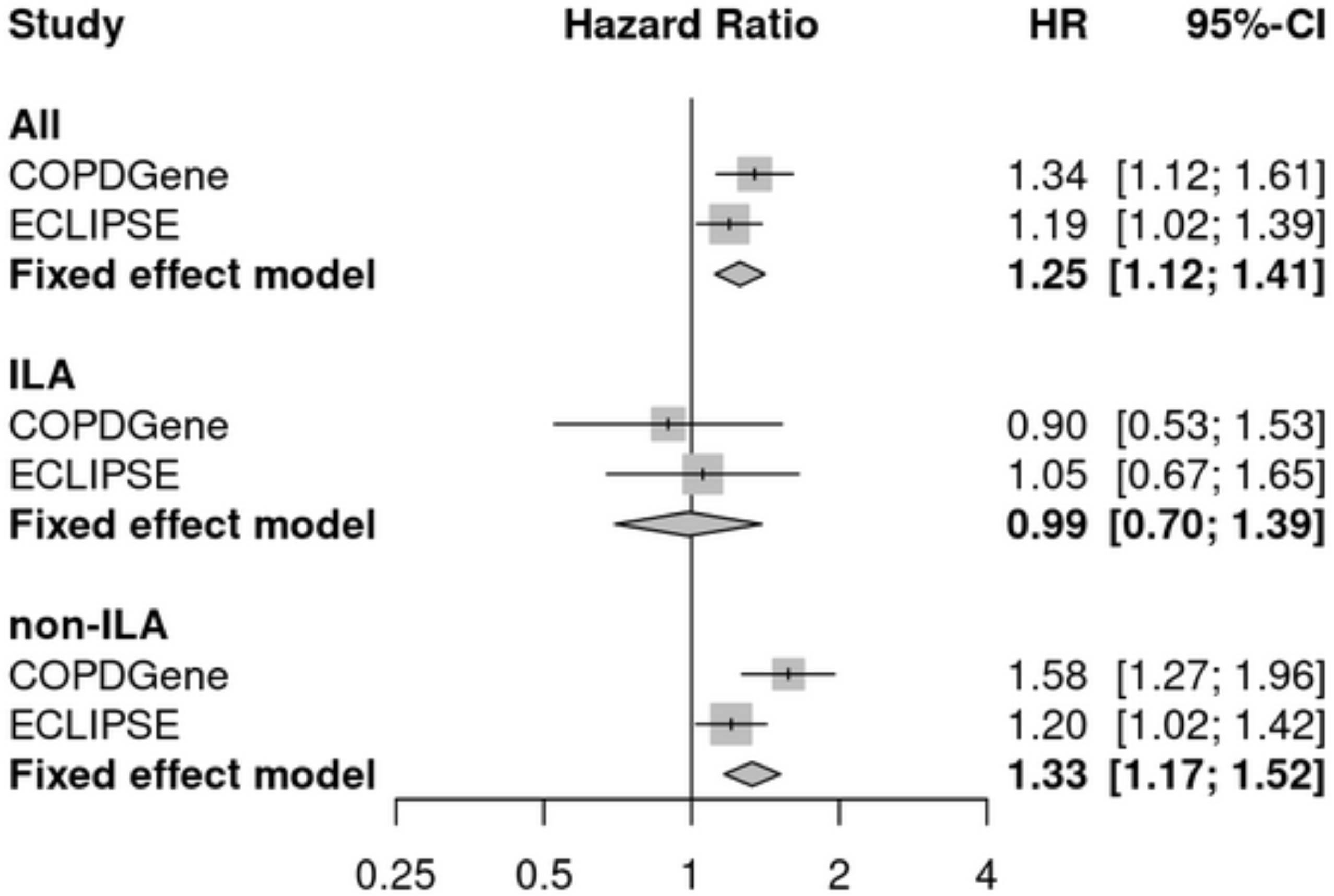
Forest plots based on time-to-death analyses of the ILA score [IPF transcripts] in the COPDGene test set (n=735) and ECLIPSE (n=571). Cox proportional hazards models were fit, adjusting for age, sex, race, body-mass index, pack-years of smoking, and current smoking. Hazard ratios are shown as per standard deviation increase in the ILA score. ILA=interstitial lung abnormalities.

We examined the ILA score [all transcripts] for association with ILA and all-cause mortality. We found that this new score was associated with ILA and all-cause mortality in the COPDGene testing set, but only with all-cause mortality in ECLIPSE (Table E6).

### Characterization of the gene signature

Having demonstrated that the genes in the IPF mortality score can be re-weighted to create an ILA score, and that this ILA score [IPF transcripts] consistently associates with ILA and time-to-death in two cohorts, we sought to understand how biological processes annotated to the IPF mortality genes relate to our outcomes. We observed that these 50 genes are highly correlated with each other and may represent coordinated biological processes. A heatmap of Pearson correlation coefficients is shown in Figure E3. In pathway enrichment analyses, the 50 IPF score genes were associated with three pathways: Butyrophilin family interactions, P2Y receptors, and immunoregulatory interactions between lymphoid and non-lymphoid cells (Table E7). The effects of varying levels of genes in the ILA score [IPF transcripts] on predictive performance for ILA are shown in the supplement (Supplementary Results; Table E8; Figure E4).

## Discussion

While there has been extensive research into gene expression changes associated with IPF^36–39^, no studies, to our knowledge, have examined gene expression profiles in ILA. ILA may progress to pulmonary fibrosis in certain instances^4,5^, and IPF and ILA have demonstrated overlapping yet distinct genetic underpinnings^10^. In this study of over 2,000 individuals with blood gene expression and ILA phenotype data, we examined the association of a blood IPF gene expression mortality score with ILA and all-cause mortality. We found that a previously described IPF score was not associated with ILA or mortality. By applying penalized regression to the IPF gene transcripts, we developed an 11-transcript ILA score. Six of these genes had discordant directions of effects compared to the IPF score, which may implicate important mechanisms regulating whether an individual develops ILA versus progressing to pulmonary fibrosis. While there may be some shared pathogenic mechanisms between ILA and IPF, the amount of discordance we observed further suggests that some of those with ILA (among populations of smokers) are likely distinct from IPF. Two ILA scores, derived from IPF score or genome-wide transcripts, were associated with all-cause mortality in both cohorts, suggesting that the transcripts relevant to ILA risk may represent general risk factors for mortality. We identified a peripheral blood signature of ILA, demonstrated overlapping and distinct gene transcripts between ILA and IPF, and lend insight into how gene expression profiles and biological pathways associated with IPF prognosis relate to ILA and all-cause mortality.

Our analyses may lend insight into biological processes relevant to ILA. We developed our risk score using LASSO, a method that reduces collinearity amongst features and optimizes prediction accuracy, but which does not necessarily choose the most biologically relevant features; thus, biological interpretation must be performed with caution. Despite this caveat, many of the genes identified as being predictive of ILA have been implicated in IPF pathogenesis, and the correlation structure amongst IPF risk score genes allowed us to perform pathway enrichment analyses and link the predictive transcripts to specific biological processes.

Five genes demonstrated concordant directions of effects between the ILA [IPF transcripts] and IPF scores *(CXCR6, IL7R, LBH, LRRC39, PLBD1)*, suggesting these genes may represent important biologic processes in promoting the progression of pulmonary fibrosis. *IL7R* had the largest absolute effect size in the ILA score [IPF transcripts] and was inversely associated with ILA. Our results are consistent with in vitro and in vivo molecular evidence demonstrating that IL-7 inhibits fibroblast TGF-ß production in pulmonary fibrosis^40^. *CXCR6* is a chemokine receptor that attracts T cells to the lungs^41^ and can promote the epithelial-mesenchymal transition in cell lines^42^, an event that may be important to the progression of pulmonary fibrosis. *LBH* is a transcription factor involved in Wnt signaling and is highly expressed in sub-populations of matrix fibroblasts from mouse lung^43^. Thus, there is already evidence that *IL7R, LBH*, and *CXCR6* are important for understanding processes that promote pulmonary fibrosis, and *LRRC39* and *PLBD1* are additional targets for future studies.

The transcripts with discordant directions of effects compared to the IPF score (*BTN3A1, GBP4, CPED1, GPR174, LPAR6, NAP1L2*) support the notion that ILA may represent a collection of disorders, some of which are distinct from IPF. This observation is consistent with genetic analyses of IPF and ILA. A genome-wide association study (GWAS) of ILA identified four genome-wide significant variants (in *FCF1P3, IPO11, HTR1E, MUC5B*); while the *MUC5B* rs35705950 promoter polymorphism is well known in IPF, the other three loci were not associated with IPF. Similarly, out 12 previously reported IPF GWAS variants, only four other variants were significantly associated with ILA (*DPP9, DSP, FAM13A, IVD*), though most of the others were consistent in effect direction. Whether the genetic variants uniquely associated with ILA indirectly regulate the discordant genes identified in the current study is unclear and requires further investigation.

The transcripts with discordant directions of effects between the ILA [IPF transcripts] and IPF scores allude to divergent biologic processes that could play a role in determining whether a person develops ILA or progresses to irreversible fibrosis. *BTN3A1* is part of the Butyrophilin immunoglobulin superfamily. Butyrophilins may play a role in facilitating interactions between adaptive and innate immune cells^44^. *GBP4* is a guanylate binding protein that facilitates second messenger signaling for interferons, and has been observed to increase in response to cytokine stimulation in IPF lungs^45^. Genetic variation in *GPR174* is associated with susceptibility to autoimmune disease, and *GPR174*-deficient mice were resistant to lipopolysaccharide-induced cytokine storm^46^. *NAP1L2* promotes histone acetylation during neuronal differentiation^47^ and other members of the same protein family have demonstrated increased expression in lung fibroblasts from IPF patients^48^. Taken together, these data suggest that immune responses to environmental/infectious insults, genetic variation, and epigenetic modifications may be important to understanding the distinct pathogenic mechanisms present in ILA that may lead to IPF.

We observed that the ILA score [IPF transcripts] was associated with all-cause mortality, but that this association was driven by non-ILA participants, albeit with small samples sizes for ILA. In single gene association analyses for the 50 IPF score genes, nearly half of all transcripts had discordant effect directions for ILA association compared to associations with all-cause mortality and in comparison to gene weights in the IPF score. Conversely, the 50 IPF gene effect directions in association with all-cause mortality (in all study participants) were highly concordant with the directions of weights in the IPF score with only three genes having discordant effect directions. Further, an ILA score trained using genome-wide transcripts in the COPDGene training data was not associated with ILA in both the COPDGene testing set and ECLIPSE but was associated with all-cause mortality in both cohorts. These data suggest that transcripts associated with ILA risk may also be risk factors for all-cause mortality. When training a gene expression model to ILA, we identified a gene set associated with all-cause mortality; however, an ILA score derived from the limited set of IPF score genes is associated with ILA and mortality. These observations suggest that the IPF score represents a mixture of transcripts relevant to all-cause mortality and the ILA-IPF axis.

*LPAR6* was the only non-HLA transcript with concordant directions of effects when tested for association with ILA and all-cause mortality and had discordant directions of effects compared to the IPF score. *LPAR6* is a lysophosphatidic acid (LPA) receptor. LPA has been shown to signal through its G-protein-coupled receptors to induce pro-inflammatory signals from stressed epithelial cells and activate TGF signaling^49^. Thus, LPAR6 could represent a shared mechanism between ILA and all-cause mortality or it could be on the causal pathway between ILA and death.

As aging is a major driver of mortality, we sought to determine whether the effects of the ILA score [IPF transcripts] on mortality represent an aging effect. In causal mediation analyses, we found that about 12% of the effect of age on mortality was mediated through the ILA score [IPF transcripts]. Thus, the 11 genes in the ILA score [IPF transcripts] appear to be important for overall mortality and partially represent aging effects on mortality. However, the majority of aging effects are not captured by the ILA score [IPF transcripts], which highlights the need for further research into the shared and divergent biological processes related to ILA, aging, and mortality.

Strengths of this study include replication in two well-characterized cohorts of smokers, cross-technology replication (both RNA-seq and microarray), and the application of causal inference analyses to examine the relationship between IPF and ILA transcriptomic risk with aging and risk of death. However, we were not able to assess the association of the ILA risk scores with the development or progression of ILA due to data availability. Our primary analysis was of genes identified in a study of IPF, which allowed an assessment of the contrasts between IPF mortality and ILA risk in a well-validated set of genes. Notably, attempting to use a larger set of genes in the blood transcriptome, including other transcripts that may be more predictive of ILA risk, was not superior. Other approaches using blood gene expression and additional -Omics data (such as methylation, proteomic, microbiome) may allow us to delve deeper into the mechanisms underlying our observed associations, as well as longitudinal studies of ILA and IPF.

In conclusion, a peripheral blood gene signature associated with IPF mortality was not associated with ILA or mortality in two well-characterized cohorts of smokers. An ILA gene expression score, derived from the genes in the IPF score, was reproducibly associated with ILA and all-cause mortality in current and former smokers. Separately, an ILA score derived from genome-wide transcripts was associated with all-cause mortality, but not ILA in validation studies. Approximately half of the genes in the ILA score [IPF transcripts] were of opposite direction in the IPF score, and genes associated with ILA may also be risk factors for mortality and partially represent aging effects on mortality. Genes identified in this study may be important candidates to further examine in the pathogenesis and progression of ILA to IPF and in mortality.

## Data Availability

All data produced in the present work are contained in the manuscript and access to raw data are available upon reasonable request to the authors

