## Supplementary Appendix for "Blood Gene Expression Risk Profiles and Interstitial Lung Abnormalities: COPDGene and ECLIPSE cohort studies"

**Table of Contents**

Funding and Acknowledgements	2

Supplementary Methods	6

Supplementary Results	7

Supplementary Tables

Table E1	8

Table E2	11

Table E3	12

Table E4	13

Table E5	14

Table E6	15

Table E7	16

Table E8	17

Supplementary Figures

Figure E1	18

Figure E2	19

Figure E3	20

Figure E4	21

References………………………………………………………………………………………..22

**Funding and Acknowledgements**

**COPDGene Phase 3**

**Grant Support and Disclaimer**

The project described was supported by Award Number U01 HL089897 and Award Number U01 HL089856 from the National Heart, Lung, and Blood Institute. The content is solely the responsibility of the authors and does not necessarily represent the official views of the National Heart, Lung, and Blood Institute or the National Institutes of Health.

**COPD Foundation Funding**

COPDGene is also supported by the COPD Foundation through contributions made to an Industry Advisory Board comprised of AstraZeneca, Boehringer-Ingelheim, Genentech, GlaxoSmithKline, Novartis, Pfizer, Siemens, and Sunovion.

**COPDGene^®^ Investigators – Core Units**

*Administrative Center*: James D. Crapo, MD (PI); Edwin K. Silverman, MD, PhD (PI); Barry J. Make, MD; Elizabeth A. Regan, MD, PhD

*Genetic Analysis Center*: Terri Beaty, PhD; Ferdouse Begum, PhD; Peter J. Castaldi, MD, MSc; Michael Cho, MD; Dawn L. DeMeo, MD, MPH; Adel R. Boueiz, MD; Marilyn G. Foreman, MD, MS; Eitan Halper-Stromberg; Lystra P. Hayden, MD, MMSc; Craig P. Hersh, MD, MPH; Jacqueline Hetmanski, MS, MPH; Brian D. Hobbs, MD; John E. Hokanson, MPH, PhD; Nan Laird, PhD; Christoph Lange, PhD; Sharon M. Lutz, PhD; Merry-Lynn McDonald, PhD; Margaret M. Parker, PhD; Dmitry Prokopenko, Ph.D; Dandi Qiao, PhD; Elizabeth A. Regan, MD, PhD; Phuwanat Sakornsakolpat, MD; Edwin K. Silverman, MD, PhD; Emily S. Wan, MD; Sungho Won, PhD

*Imaging Center*: Juan Pablo Centeno; Jean-Paul Charbonnier, PhD; Harvey O. Coxson, PhD; Craig J. Galban, PhD; MeiLan K. Han, MD, MS; Eric A. Hoffman, Stephen Humphries, PhD; Francine L. Jacobson, MD, MPH; Philip F. Judy, PhD; Ella A. Kazerooni, MD; Alex Kluiber; David A. Lynch, MB; Pietro Nardelli, PhD; John D. Newell, Jr., MD; Aleena Notary; Andrea Oh, MD; Elizabeth A. Regan, MD, PhD; James C. Ross, PhD; Raul San Jose Estepar, PhD; Joyce Schroeder, MD; Jered Sieren; Berend C. Stoel, PhD; Juerg Tschirren, PhD; Edwin Van Beek, MD, PhD; Bram van Ginneken, PhD; Eva van Rikxoort, PhD; Gonzalo Vegas Sanchez-Ferrero, PhD; Lucas Veitel; George R. Washko, MD; Carla G. Wilson, MS;

*PFT QA Center, Salt Lake City, UT*: Robert Jensen, PhD

*Data Coordinating Center and Biostatistics*, *National Jewish Health, Denver, CO*: Douglas Everett, PhD; Jim Crooks, PhD; Katherine Pratte, PhD; Matt Strand, PhD; Carla G. Wilson, MS

*Epidemiology Core*, *University of Colorado Anschutz Medical Campus, Aurora, CO*: John E. Hokanson, MPH, PhD; Gregory Kinney, MPH, PhD; Sharon M. Lutz, PhD; Kendra A. Young, PhD

*Mortality Adjudication Core:* Surya P. Bhatt, MD; Jessica Bon, MD; Alejandro A. Diaz, MD, MPH; MeiLan K. Han, MD, MS; Barry Make, MD; Susan Murray, ScD; Elizabeth Regan, MD; Xavier Soler, MD; Carla G. Wilson, MS

*Biomarker Core*: Russell P. Bowler, MD, PhD; Katerina Kechris, PhD; Farnoush Banaei-Kashani, Ph.D    **COPDGene^®^ Investigators – Clinical Centers**

*Ann Arbor VA:* Jeffrey L. Curtis, MD; Perry G. Pernicano, MD

*Baylor College of Medicine, Houston, TX*: Nicola Hanania, MD, MS; Mustafa Atik, MD; Aladin Boriek, PhD; Kalpatha Guntupalli, MD; Elizabeth Guy, MD; Amit Parulekar, MD;

*Brigham and Women’s Hospital, Boston, MA*: Dawn L. DeMeo, MD, MPH; Alejandro A. Diaz, MD, MPH; Lystra P. Hayden, MD; Brian D. Hobbs, MD; Craig Hersh, MD, MPH; Francine L. Jacobson, MD, MPH; George Washko, MD

*Columbia University, New York, NY*: R. Graham Barr, MD, DrPH; John Austin, MD; Belinda D’Souza, MD; Byron Thomashow, MD

*Duke University Medical Center, Durham, NC*: Neil MacIntyre, Jr., MD; H. Page McAdams, MD; Lacey Washington, MD

*Grady Memorial Hospital, Atlanta, GA: Eric Flenaugh, MD; Silanth Terpenning, MD*

*HealthPartners Research Institute, Minneapolis, MN*: Charlene McEvoy, MD, MPH; Joseph Tashjian, MD

*Johns Hopkins University, Baltimore, MD*: Robert Wise, MD; Robert Brown, MD; Nadia N. Hansel, MD, MPH; Karen Horton, MD; Allison Lambert, MD, MHS; Nirupama Putcha, MD, MHS

*Lundquist Institute for Biomedical Innovationat Harbor UCLA Medical Center, Torrance, CA*: Richard Casaburi, PhD, MD; Alessandra Adami, PhD; Matthew Budoff, MD; Hans Fischer, MD; Janos Porszasz, MD, PhD; Harry Rossiter, PhD; William Stringer, MD

*Michael E. DeBakey VAMC, Houston*, *TX*: Amir Sharafkhaneh, MD, PhD; Charlie Lan, DO

*Minneapolis VA:* Christine Wendt, MD; Brian Bell, MD; Ken M. Kunisaki, MD, MS

*National Jewish Health, Denver, CO*: Russell Bowler, MD, PhD; David A. Lynch, MB

*Reliant Medical Group, Worcester, MA*: Richard Rosiello, MD; David Pace, MD

*Temple University, Philadelphia, PA:* Gerard Criner, MD; David Ciccolella, MD; Francis Cordova, MD; Chandra Dass, MD; Gilbert D’Alonzo, DO; Parag Desai, MD; Michael Jacobs, PharmD; Steven Kelsen, MD, PhD; Victor Kim, MD; A. James Mamary, MD; Nathaniel Marchetti, DO; Aditi Satti, MD; Kartik Shenoy, MD; Robert M. Steiner, MD; Alex Swift, MD; Irene Swift, MD; Maria Elena Vega-Sanchez, MD

*University of Alabama, Birmingham, AL:* Mark Dransfield, MD; William Bailey, MD; Surya P. Bhatt, MD; Anand Iyer, MD; Hrudaya Nath, MD; J. Michael Wells, MD

*University of California, San Diego, CA*: Douglas Conrad, MD; Xavier Soler, MD, PhD; Andrew Yen, MD

*University of Iowa, Iowa City, IA*: Alejandro P. Comellas, MD; Karin F. Hoth, PhD; John Newell, Jr., MD; Brad Thompson, MD

*University of Michigan, Ann Arbor, MI:* MeiLan K. Han, MD MS; Ella Kazerooni, MD MS; Wassim Labaki, MD MS; Craig Galban, PhD; Dharshan Vummidi, MD

*University of Minnesota, Minneapolis, MN*: Joanne Billings, MD; Abbie Begnaud, MD; Tadashi Allen, MD

*University of Pittsburgh, Pittsburgh, PA*: Frank Sciurba, MD; Jessica Bon, MD; Divay Chandra, MD, MSc; Joel Weissfeld, MD, MPH

*University of Texas Health, San Antonio, San Antonio, TX*: Antonio Anzueto, MD; Sandra Adams, MD; Diego Maselli-Caceres, MD; Mario E. Ruiz, MD; Harjinder Singh

**Supplementary Methods**

For the ILA score [IPF transcripts], we also sought to determine how the number of included genes affected the predictive performance. We ranked ILA score [IPF transcripts] gene transcripts based on absolute values of effect sizes. We then constructed logistic regression models with ILA as the outcome, sequentially added transcripts from the smallest to the largest effect size and performed area-under-the-receiver-operator-characteristic-curve (AUC) analyses.

**Supplementary Results**

To understand the relative contributions of transcripts in the ILA score [IPF transcripts] to ILA, we ranked the 11 transcripts based on absolute values of effect sizes (Table E6) from the LASSO regression. *IL7R* had the largest effect size, while *NAPIL2* had the smallest effect size. Going from the smallest to the largest absolute value of effect sizes in the ILA score [IPF transcripts], we sequentially added transcripts to logistic regression models of ILA and performed AUC analyses (Figure E4). We observed that the optimal AUC was obtained with 11 transcripts, yet with overlapping confidence intervals compared to models with smaller numbers of transcripts.

**Supplementary Tables**

Table E1: Associations of individual gene transcripts with ILA and all-cause mortality were performed with logistic regression. ILA beta coefficients were compared to the direction of expression in the IPF risk score and to the directions of effects in all-cause mortality associations. The directions of effects of transcripts on ILA were discordant compared to the directions of effects on all-cause mortality (24 transcripts) and in the IPF score (23 transcripts). The directions of effects of transcripts on all-cause mortality were only discordant with the mortality score for three transcripts (*HLA-DP1*, *HLA-DP2*, and *LPAR6*). Logistic regression models were adjusted for age, sex, race, body-mass index, pack-years of smoking, and current smoking status. *: p-value < 0.05. ** = present in ILA score.

| *HGNC symbol* | *beta (95% CI)* | *p* | *Direction in IPF score* | *ILA vs. IPF score* |  | *ILA vs. Mortality Association* |
| --- | --- | --- | --- | --- | --- | --- |
| *ARHGAP5* | -0.0077 (-0.21-0.2) | 0.94 | - | Concordant down |  | Concordant down |
| *ARL4C* | -0.12 (-0.32-0.083) | 0.25 | - | Concordant down |  | Concordant down |
| *BIRC3* | -0.0057 (-0.21-0.2) | 0.96 | - | Concordant down |  | Concordant down |
| *BTN3A1* | 0.23 (0.0072-0.46) | 0.043* | - | Discordant (ILA score up) | ** | Discordant (ILA score up) |
| *BTN3A2* | 0.012 (-0.2-0.22) | 0.91 | - | Discordant (ILA score up) |  | Discordant (ILA score up) |
| *BTN3A3* | 0.14 (-0.075-0.36) | 0.2 | - | Discordant (ILA score up) |  | Discordant (ILA score up) |
| *CAMK2D* | 0.062 (-0.15-0.28) | 0.57 | - | Discordant (ILA score up) |  | Discordant (ILA score up) |
| *CD2* | -0.019 (-0.23-0.19) | 0.86 | - | Concordant down |  | Concordant down |
| *CD28* | -0.077 (-0.29-0.13) | 0.47 | - | Concordant down |  | Concordant down |
| *CD47* | 0.077 (-0.13-0.29) | 0.47 | - | Discordant (ILA score up) |  | Discordant (ILA score up) |
| *CD96* | -0.056 (-0.27-0.16) | 0.61 | - | Concordant down |  | Concordant down |
| *CNOT6L* | 0.017 (-0.19-0.22) | 0.87 | - | Discordant (ILA score up) |  | Discordant (ILA score up) |
| *CPED1* | 0.087 (-0.13-0.3) | 0.43 | - | Discordant (ILA score up) | ** | Discordant (ILA score up) |
| *CXCR6* | -0.085 (-0.29-0.12) | 0.41 | - | Concordant down | ** | Concordant down |
| *DOCK10* | 0.0041 (-0.22-0.22) | 0.97 | - | Discordant (ILA score up) |  | Discordant (ILA score up) |
| *DYNC2LI1* | 0.076 (-0.13-0.28) | 0.47 | - | Discordant (ILA score up) |  | Discordant (ILA score up) |
| *ETS1* | -0.11 (-0.33-0.11) | 0.32 | - | Concordant down |  | Concordant down |
| *FLT3* | -0.11 (-0.33-0.1) | 0.3 | + | Discordant (ILA score down) |  | Discordant (ILA score down) |
| *GBP4* | 0.28 (0.047-0.51) | 0.018* | - | Discordant (ILA score up) | ** | Discordant (ILA score up) |
| *GPR174* | 0.12 (-0.098-0.34) | 0.28 | - | Discordant (ILA score up) | ** | Discordant (ILA score up) |
| *GPR183* | -0.11 (-0.31-0.093) | 0.29 | - | Concordant down |  | Concordant down |
| *HLA-DPA1* | -0.0059 (-0.21-0.2) | 0.96 | - | Concordant down |  | Discordant (ILA score down) |
| *HLA-DPB1* | -0.038 (-0.25-0.17) | 0.72 | - | Concordant down |  | Discordant (ILA score down) |
| *HP* | -0.026 (-0.26-0.21) | 0.83 | + | Discordant (ILA score down) |  | Discordant (ILA score down) |
| *ICOS* | 0.037 (-0.18-0.25) | 0.73 | - | Discordant (ILA score up) |  | Discordant (ILA score up) |
| *IL1R2* | -0.018 (-0.22-0.18) | 0.86 | + | Discordant (ILA score down) |  | Discordant (ILA score down) |
| *IL7R* | -0.19 (-0.41-0.023) | 0.08 | - | Concordant down | ** | Concordant down |
| *ITK* | -0.053 (-0.27-0.16) | 0.62 | - | Concordant down |  | Concordant down |
| *KLF12* | -0.042 (-0.25-0.17) | 0.69 | - | Concordant down |  | Concordant down |
| *LARP4* | 0.081 (-0.13-0.29) | 0.45 | - | Discordant (ILA score up) |  | Discordant (ILA score up) |
| *LBH* | -0.29 (-0.5--0.079) | 0.0069* | - | Concordant down | ** | Concordant down |
| *LCK* | -0.14 (-0.34-0.067) | 0.19 | - | Concordant down |  | Concordant down |
| *LPAR6* | 0.14 (-0.093-0.36) | 0.25 | - | Discordant (ILA score up) | ** | Concordant up |
| *LRRC39* | -0.034 (-0.24-0.17) | 0.74 | - | Concordant down | ** | Concordant down |
| *MCEMP1* | 0.11 (-0.11-0.34) | 0.32 | + | Concordant up |  | Concordant up |
| *METTL8* | -0.00097 (-0.21-0.21) | 0.99 | - | Concordant down |  | Concordant down |
| *MORC4* | -0.087 (-0.3-0.12) | 0.42 | - | Concordant down |  | Concordant down |
| *NAP1L2* | -0.04 (-0.24-0.16) | 0.69 | - | Concordant down | ** | Concordant down |
| *NUP43* | -0.042 (-0.25-0.16) | 0.69 | - | Concordant down |  | Concordant down |
| *P2RY10* | -0.062 (-0.28-0.16) | 0.58 | - | Concordant down |  | Concordant down |
| *PLBD1* | 0.18 (-0.051-0.42) | 0.12 | + | Concordant up | ** | Concordant up |
| *S100A12* | 0.092 (-0.14-0.33) | 0.44 | + | Concordant up |  | Concordant up |
| *S1PR1* | -0.17 (-0.38-0.041) | 0.11 | - | Concordant down |  | Concordant down |
| *SH2D1A* | 0.041 (-0.18-0.26) | 0.72 | - | Discordant (ILA score up) |  | Discordant (ILA score up) |
| *SLAMF7* | 0.074 (-0.14-0.29) | 0.5 | - | Discordant (ILA score up) |  | Discordant (ILA score up) |
| *STAT4* | -0.084 (-0.29-0.12) | 0.42 | - | Concordant down |  | Concordant down |
| *TC2N* | 0.022 (-0.19-0.23) | 0.84 | - | Discordant (ILA score up) |  | Discordant (ILA score up) |
| *TPST1* | -0.17 (-0.39-0.054) | 0.14 | + | Discordant (ILA score down) |  | Discordant (ILA score down) |
| *TRAT1* | 0.0066 (-0.19-0.2) | 0.95 | - | Discordant (ILA score up) |  | Discordant (ILA score up) |
| *UTRN* | 0.11 (-0.11-0.33) | 0.35 | - | Discordant (ILA score up) |  | Discordant (ILA score up) |

Table E2: The IPF risk score was re-weighted to create an ILA score [IPF transcripts]. New weights are shown. For the ILA score, the intercept is -1.687029, and 11 transcripts were selected. Note that two transcripts were not available in both of our datasets (*C2ORF27A, SNHG1*). HGNC=HUGO Gene Nomenclature Committee. ILA=interstitial lung abnormalities. + = transcript expression up; - = transcript expression down. Transcripts are arranged in alphabetical order based on HGNC symbol.

| *HGNC symbol* | *Ensemble ID* | *ILA score weight* | *Direction in IPF score* | *ILA vs. IPF score* |
| --- | --- | --- | --- | --- |
| *BTN3A1* | ENSG00000026950 | 0.167 | - | Discordant (ILA score up) |
| *CPED1* | ENSG00000106034 | 0.0289 | - | Discordant (ILA score up) |
| *CXCR6* | ENSG00000172215 | -0.0843 | - | Concordant down |
| *GBP4* | ENSG00000162654 | 0.05 | - | Discordant (ILA score up) |
| *GPR174* | ENSG00000147138 | 0.175 | - | Discordant (ILA score up) |
| *IL7R* | ENSG00000168685 | -0.49 | - | Concordant down |
| *LBH* | ENSG00000213626 | -0.18 | - | Concordant down |
| *LPAR6* | ENSG00000139679 | 0.107 | - | Discordant (ILA score up) |
| *LRRC39* | ENSG00000122477 | -0.0239 | - | Concordant down |
| *NAP1L2* | ENSG00000186462 | 0.000859 | - | Discordant (ILA score up) |
| *PLBD1* | ENSG00000121316 | 0.114 | + | Concordant up |

Table E3: A new ILA score was derived from genome-wide transcripts (ILA score [all transcripts]). The resulting score had 25 transcripts, none of which were in the IPF score. The intercept for this model was -1.6991577.

| *HGNC symbol* | *weight* |
| --- | --- |
| *FLVCR1* | 0.178332805 |
| *SLC8A1* | 0.037822338 |
| *EML6* | -0.043367016 |
| *PDGFC* | 0.01925094 |
| *TMEM144* | 0.035076472 |
| *SLC26A8* | 0.067644632 |
| *BACH2* | -0.047060442 |
| *MARCKS* | 0.025317428 |
| *MEST* | -0.007792345 |
| *PLAG1* | -0.219186046 |
| *TG* | 0.049046419 |
| *MPZL2* | 0.019174067 |
| *JAM3* | -0.047222304 |
| *AKR1E2* | -0.109435648 |
| *MYOF* | 0.042876457 |
| *TCF7L2* | 0.046696665 |
| *DHRS12* | 0.076456282 |
| *EGLN3* | -0.045672989 |
| *TTC9* | -0.046100963 |
| *TDRD9* | 0.003350668 |
| *APBA2* | -0.070753249 |
| *THBS1* | -0.075916443 |
| *SREBF1* | -0.011136454 |
| *TSHZ3* | 0.023575896 |

Table E4: Odds ratios of the IPF and ILA [IPF transcripts] risk scores with ILA in the COPDGene test set (n=735) and ECLIPSE (n=571), adjusted for white cell counts. Logistic regression models were adjusted for age, sex, race, body-mass index, pack-years of smoking, current smoking status, and white blood cell counts. Inverse variance fixed-effects meta-analyses of the adjusted estimates were performed. ILA=interstitial lung abnormalities. IPF=idiopathic pulmonary fibrosis. The Bonferroni-adjusted p-value threshold is 0.05/2 risk scores/2 cohorts = 0.0125.

| *Score* | COPDGene | | | | ECLIPSE | | | | Combined (Meta-analyses) | |
| --- | --- | --- | --- | --- | --- | --- | --- | --- | --- | --- |
|  | Unadjusted |  | Adjusted |  | Unadjusted |  | Adjusted |  | Adjusted |  |
|  | *OR (95% CI)* | *p* | *OR (95% CI)* | *p* | *OR (95% CI)* | *p* | *OR (95% CI)* | *p* | *OR (95% CI)* | *p* |
| IPF score | 1.8 (1.1-2.8) | 0.018 | 1.7 (1.1-2.8) | 0.023 | 0.64 (0.22-1.9) | 0.41 | 0.63 (0.22-1.8) | 0.4 | 1.5 (0.95 - 2.3) | 0.083 |
| ILA score [IPF transcripts] | 1.4 (1.2-1.7) | 6.70E-05 | 1.3 (1.1-1.6) | 0.0019 | 1.5 (1.2-2) | 0.001 | 1.4 (1.1-1.9) | 0.011 | 1.4 (1.2 - 1.6) | 6.80E-05 |

Table E5: Association of the IPF and ILA [IPF transcripts] scores with time-to-death in the COPDGene test set (n=735) and ECLIPSE (n=571), adjusted for white blood cell counts. Multivariable Cox regression models were adjusted for age, sex, race, body-mass index, pack-years of smoking, current smoking status, and white blood cell counts. ILA = interstitial lung abnormalities.

| *Score* | *Cohort* | *Stratum* | *Unadjusted* | | *Adjusted* | |
| --- | --- | --- | --- | --- | --- | --- |
|  |  |  | HR (95% CI) | p | HR (95% CI) | p |
| IPF score | COPDGene test set | All | 1.3 (0.71-2.3) | 0.41 | 1.3 (0.7-2.3) | 0.44 |
|  |  | ILA | 1.3 (0.47-3.8) | 0.6 | 1.2 (0.44-3.6) | 0.68 |
|  |  | non-ILA | 1.2 (0.57-2.4) | 0.67 | 1.2 (0.58-2.4) | 0.63 |
|  | ECLIPSE | All | 1.3 (0.77-2.1) | 0.36 | 1.2 (0.75-2) | 0.4 |
|  |  | ILA | 2.2 (0.49-10) | 0.3 | 2.6 (0.54-13) | 0.23 |
|  |  | non-ILA | 1.2 (0.72-2.1) | 0.47 | 1.2 (0.7-2) | 0.54 |
| ILA score [IPF transcripts] | COPDGene test set | All | 1.4 (1.2-1.7) | 3.60E-05 | 1.3 (1.1-1.6) | 0.0012 |
|  |  | ILA | 1 (0.64-1.6) | 0.99 | 0.9 (0.53-1.5) | 0.69 |
|  |  | non-ILA | 1.6 (1.3-2) | 2.50E-06 | 1.6 (1.3-2) | 4.00E-05 |
|  | ECLIPSE | All | 1.3 (1.1-1.4) | 0.003 | 1.2 (1-1.4) | 0.023 |
|  |  | ILA | 1.2 (0.78-1.7) | 0.47 | 1.1 (0.67-1.7) | 0.82 |
|  |  | non-ILA | 1.3 (1.1-1.5) | 0.006 | 1.2 (1-1.4) | 0.027 |

Table E6: A new ILA score developed using all available transcripts (ILA score [all transcripts]) was tested for association with ILA and all-cause mortality in the COPDGene testing set and ECLIPSE. Logistic regression models were performed for ILA, and Cox regression for time-to-death. All models were adjusted for age, sex, race, body-mass index, pack-years of smoking, and current smoking status. ILA = Interstitial lung abnormalities.

| *cohort* | *outcome* | *score* | Unadjusted | | Adjusted | |
| --- | --- | --- | --- | --- | --- | --- |
|  |  |  | *OR (95% CI)* | *p* | *OR (95% CI)* | *p* |
| COPDGene Test | ILA | ILA score [all transcripts] | 1.6 (1.3 to 1.9) | 1.00E-05 | 1.4 (1.1 to 1.7) | 0.0054 |
| COPDGene Test | Time-to-death | ILA score [all transcripts] | 1.5 (1.1 to 1.9) | 0.0019 | 1.3 (1 to 1.7) | 0.037 |
| ECLIPSE | ILA | ILA score [all transcripts] | 1.3 (0.94 to 1.9) | 0.1 | 1.1 (0.79 to 1.6) | 0.47 |
| ECLIPSE | Time-to-death | ILA score [all transcripts] | 1.4 (1.1 to 1.6) | 0.00093 | 1.3 (1 to 1.5) | 0.023 |

Table E7: Top 10 Reactome pathways^1^ associated with the 52-gene signature.

| *Reactome ID* | *Pathway Description* | *p-values* | *Bonferroni-adjusted p-values* | *No. of pathways examined* |
| --- | --- | --- | --- | --- |
| R-HSA-8851680 | Butyrophilin (BTN) family interactions | 5.46E-18 | 5.46E-15 | 1001 |
| R-HSA-417957 | P2Y receptors | 2.30E-08 | 2.30E-05 | 1001 |
| R-HSA-198933 | Immunoregulatory interactions between a Lymphoid and a non-Lymphoid cell | 1.62E-06 | 0.001621 | 1001 |
| R-HSA-2168880 | Scavenging of heme from plasma | 0.003503 | 1 | 285 |
| R-HSA-373076 | Class A/1 (Rhodopsin-like receptors) | 0.0042 | 1 | 238 |
| R-HSA-2559585 | Oncogene Induced Senescence | 0.004462 | 1 | 224 |
| R-HSA-419408 | Lysosphingolipid and LPA receptors | 0.006925 | 1 | 144 |
| R-HSA-9020702 | Interleukin-1 signaling | 0.01115 | 1 | 90 |
| R-HSA-6804115 | TP53 regulates transcription of additional cell cycle genes whose exact role in the p53 pathway remain uncertain | 0.01265 | 1 | 79 |
| R-HSA-202733 | Cell surface interactions at the vascular wall | 0.01418 | 1 | 71 |

Table E8: ILA score [IPF transcripts] transcripts ranked by absolute value of effect size. ILA = interstitial lung abnormalities. HGNC = HUGO Gene Nomenclature Committee.

| *HGNC symbol* | *Ensemble ID* | *beta* |
| --- | --- | --- |
| *IL7R* | ENSG00000168685 | -0.49 |
| *LBH* | ENSG00000213626 | -0.18 |
| *GPR174* | ENSG00000147138 | 0.18 |
| *BTN3A1* | ENSG00000026950 | 0.17 |
| *PLBD1* | ENSG00000121316 | 0.11 |
| *LPAR6* | ENSG00000139679 | 0.11 |
| *CXCR6* | ENSG00000172215 | -0.084 |
| *GBP4* | ENSG00000162654 | 0.05 |
| *CPED1* | ENSG00000106034 | 0.029 |
| *LRRC39* | ENSG00000122477 | -0.024 |
| *NAP1L2* | ENSG00000186462 | 0.00086 |

**Supplementary Figures**

Figure E1: Distributions of standardized ILA scores [IPF transcripts] in COPDGene (median -0.21 [interquartile range: -0.67 – 0.38]) and ECLIPSE (median -0.22 [interquartile range: -0.70 – 0.48]).

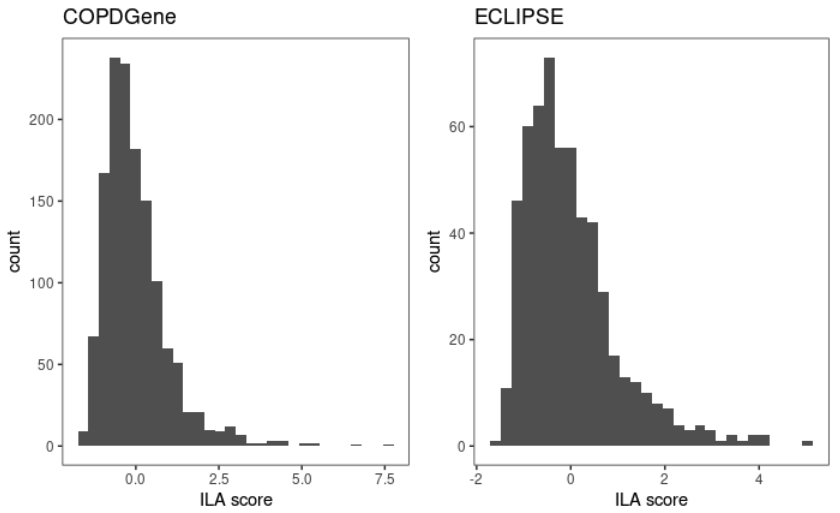

Figure E2: Histograms of the predicted probabilities for individuals in the COPDGene test set (n=735) for the IPF score for being at high risk of mortality (*blue*) and the ILA score [IPF transcripts] for ILA (*red*).

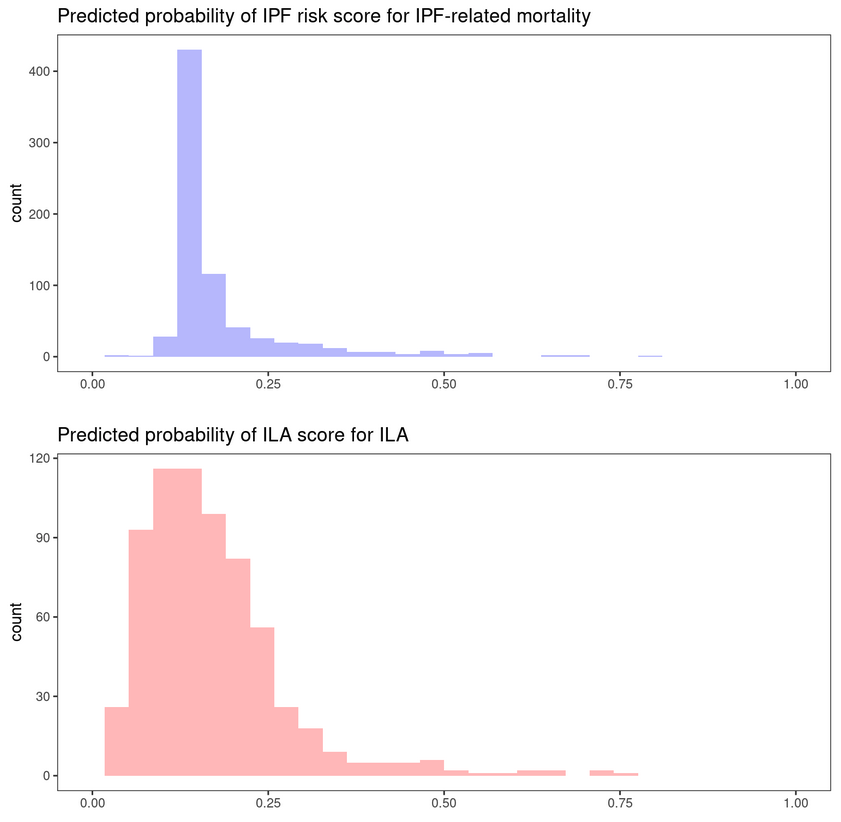

Figure E3: Heatmap of Pearson correlation coefficients demonstrating the correlation of the 50 genes with each other in the COPDGene training set (n=734).

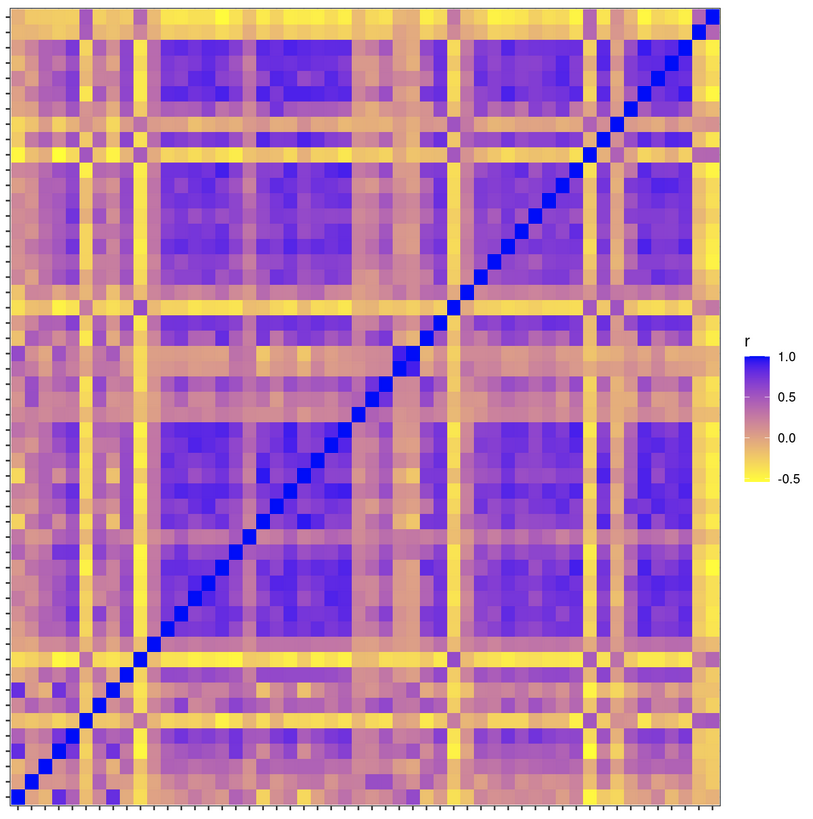

Figure E4: Area-under-the-receiver-operator-characteristic-curve (AUC) analyses of logistic regression models with ILA as the outcome, in which transcripts in the ILA score [IPF transcripts] were sequentially added to the model. Transcripts were ranked according to absolute values of effect sizes in the ILA score [IPF transcripts], and sequentially added from smallest to largest effect size. The shaded area represents DeLong 95% confidence bands.

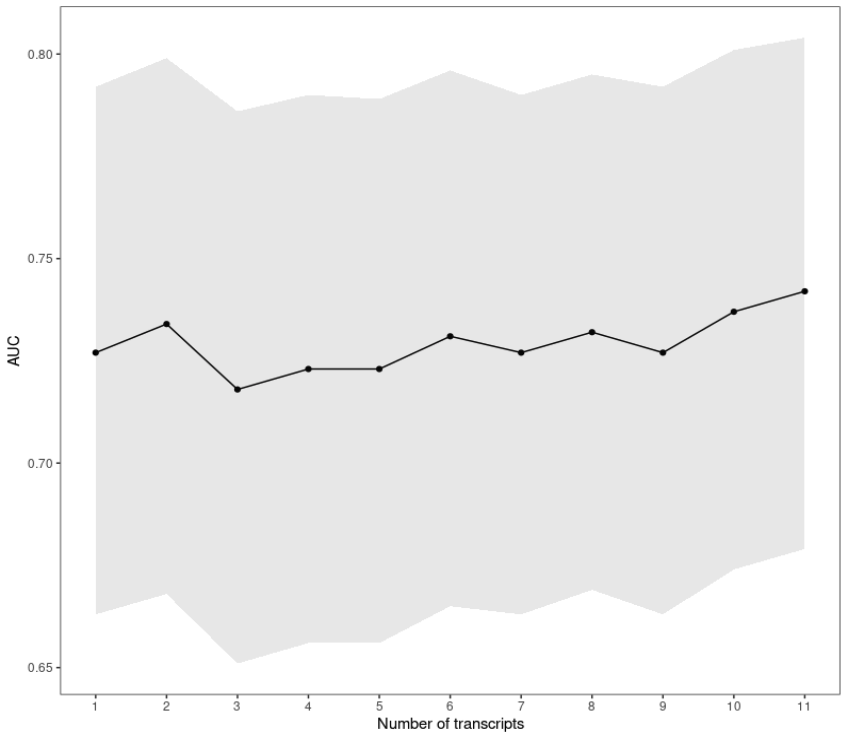
